# Associations of rope-skipping and endurance running performance with myopia and spherical equivalent refraction in children and adolescents: a cross-sectional study

**DOI:** 10.64898/2026.10.20.26365985

**Authors:** Xin Li, Aichun Li, Junlai Zhou

## Abstract

Objective To examine nonlinear associations and reference thresholds between physical fitness performance and refractive outcomes in children and adolescents. Methods We surveyed 17235 students aged 6–18 years from 35 schools in seven counties or cities in Hainan Province in December 2023. Cycloplegic autorefraction was used, and spherical equivalent refraction averaged between the two eyes (SE_mean_) of ≤−0.50 diopters was classified as myopia. Models adjusted for age, sex, ethnicity, urban–rural residence, near-work duration, outdoor activity time, parental myopia history, BMI, and a centered BMI-squared term, with school-clustered robust standard errors. Waist-to-height ratio was examined in sensitivity analyses. Results The overall myopia prevalence was 64.43%. Among students aged 6–12 years, rope skipping showed a nonlinear association with refractive outcomes, with a reference threshold of 66 skips/min. Below this threshold, each additional 10 skips/min was associated with lower odds of myopia (OR 0.949, 95% CI 0.919–0.980; P=0.001) and a 0.034-D shift toward more hyperopic SE_mean_ (95% CI 0.016– 0.052; P<0.001). Among students aged 16–18 years, endurance performance showed no clear overall nonlinearity; however, beyond 307 s, each additional 10 s was associated with higher odds of myopia (OR 1.255, 95% CI 1.030–1.531; P=0.025) and a 0.098-D shift toward myopia in SE_mean_ (95% CI −0.171 to −0.026; P=0.007). Sensitivity estimates were directionally consistent. Conclusions Fitness indicators showed range-specific associations with refractive outcomes. The two thresholds should be regarded as exploratory values for school-level risk stratification rather than causal or diagnostic cutoffs. Missing direct body-composition measures and the cross-sectional design limit interpretation of independent effects.

## Introduction

Myopia is a major public-health concern because its high prevalence is compounded by the cumulative risk of progression and later visual impairment. Projections indicate that more than 84% of children and adolescents in China may have myopia by 2050 ^[1,2]^. Evidence from epidemiological studies indicates that myopia reflects the combined effects of genetic susceptibility and environmental exposure, with environmental factors being particularly modifiable during childhood and adolescence ^[3]^. Additional time spent outdoors at school has been associated with lower myopia incidence and less myopic refractive change^[4–8]^. School schedules, recess, physical-education classes, and outdoor activities may therefore shape students’ exposure to these modifiable factors.

Most previous work has focused on outdoor activity time, which is difficult to measure precisely, whereas the relationship between routinely collected school fitness measures and refractive status has received less attention. In particular, the possibility of nonlinear associations has rarely been examined. Using screening and physical-fitness data from Han and Li students in Hainan Province, we examined age-related and population differences in myopia, evaluated nonlinear associations between rope-skipping or endurance performance and refractive outcomes, and explored whether interpretable threshold ranges could support school-level risk stratification and targeted allocation of prevention resources.

## Materials and methods

### Study population and eligibility criteria

In December 2023, we used convenience sampling to recruit students from 35 primary and secondary schools in seven counties or cities, including Haikou, Baoting, and Changjiang, Hainan Province. Physical-fitness measures were collected by trained graduate students in physical education, and refractive status was measured by professional optometrists using cycloplegic autorefraction. The survey dataset contained 18726 records. After restricting age to 6–18 years, 17336 records remained; after excluding records with unclassifiable ethnicity, 17235 remained. All students in the primary analytic sample had complete bilateral refractive measurements. The rope-skipping subgroup included students aged 6–12 years with a non-missing value greater than zero and BMI between 10 and 35 kg/m^2^ (n=8825). The endurance subgroup included students aged 16–18 years with a non-missing sex-specific 800-m or 1,000-m running time and BMI between 12 and 40 kg/m^2^ (n=3559). Written informed consent was obtained from the students and their guardians, and permission was obtained from participating schools. The study was approved by the Social Sciences Ethics Committee of Hainan Normal University (HNU-260407003) and was reported with reference to the STROBE statement^[9]^.

#### Outcome definition

Following the Technical Guidelines for the Prevention and Control of Myopia in Children and Adolescents issued by the National Health Commission of China^[10]^, refraction was measured by cycloplegic autorefraction. SE_mean_ was calculated as the mean of the right- and left-eye spherical equivalent refractions. SE_mean_≤−0.50 D was classified as myopia for the epidemiological analysis. We analyzed both the binary myopia outcome and continuous SE_mean_ to reduce information loss from reliance on a single threshold; findings were treated as relatively robust only when the directions of the two outcomes were consistent.

#### Exposure definitions

Rope skipping was defined as the number of skips completed in 1 min (skips/min). Endurance time was defined as the 800-m running time for girls and the 1000-m running time for boys, recorded in seconds.

#### Covariates and potential confounders

The basic covariates were age (continuous, years), sex, ethnicity, and urban–rural residence. Near-work duration (hours/day) and outdoor activity time (hours/day) were treated as continuous variables. Parental myopia history was coded as 0 (neither parent), 1 (one parent), or 2 (both parents), with 0 as the reference category. BMI was calculated as weight (kg)/height (m)^2^. To allow for a nonlinear association between BMI and physical performance, the primary models included centered BMI and its squared term. The dataset did not contain direct measurements of body-fat percentage, skeletal-muscle mass, or muscle quality; BMI was therefore not interpreted as a body-composition measure. Waist-to-height ratio (WHtR) was added in sensitivity analyses as a proxy for adiposity distribution.

### Statistical analysis

Analyses used two-sided tests with α=0.05. Categorical variables are presented as n (%), and continuous variables as means (standard deviations). Myopia was analyzed using a binomial logistic model, and SE_mean_ using a Gaussian model with an identity link. All primary models adjusted for age, sex, ethnicity, urban–rural residence, near-work duration, outdoor activity time, parental myopia history, BMI, and centered BMI squared. Robust standard errors were clustered by school.

#### Nonlinearity and model selection

We first fitted a linear exposure model and then fitted restricted cubic spline models for rope skipping or endurance time, with knots at the 5th, 35th, 65th, and 95th percentiles of the exposure distribution. Improvement over the linear model was assessed using the AIC difference and a joint test of nonlinearity. BMI nonlinearity was represented by the centered BMI-squared term.

#### Threshold and segmented effects

For each exposure, we constructed a single-breakpoint hinge term, max(X−c, 0). Candidate thresholds were searched over the 10th–90th exposure percentiles, and both myopia and SE_mean_ were evaluated. When thresholds differed between outcomes, the reference threshold was selected using the AIC minimum from the continuous SE_mean_ model, directional consistency of the binary outcome, and interpretability. The retained thresholds were 66 skips/min for rope skipping and 307 s for endurance time; both were treated as exploratory reference values. We report slopes before and after the threshold, 95% CIs, and tests of slope change. Effects were scaled per 10 additional skips/min and per 10-s slower endurance time.

#### Sensitivity analysis

We added WHtR to the primary models and examined whether the threshold estimates changed. This analysis was used to assess directional stability; WHtR is not equivalent to body-fat percentage or muscle mass.

## Results

### Sample composition and overall myopia prevalence

As shown in Table 1, the primary sample included 17235 students aged 6–18 years. Of these, 11104 had myopia, corresponding to a prevalence of 64.43%; the overall mean SE_mean_ was −1.41 (SD 1.84) D. Compared with students without myopia, students with myopia were older, had higher BMI and longer near-work duration, and reported less outdoor activity. The proportions of girls, Han students, and urban students were also higher in the myopia group (all P<0.001). The distribution of parental myopia history did not differ significantly between groups, but this variable was retained in the models as an important prespecified confounder. Parental myopia history was coded as 0, 1, or 2 for neither, one, or both parents, respectively; the reported P value is the overall group-comparison test.

**Table 1.** Baseline characteristics of the study population.

| Variable | Overall (n=17,235) | Myopia (n=11,104) | No myopia (n=6,131) | P value |
| --- | --- | --- | --- | --- |
| Age, years, mean (SD) | 11.83 (3.64) | 12.87 (3.38) | 9.95 (3.33) | <0.001 |
| Girls, n (%) | 8,465 (49.12) | 5,667 (51.04) | 2,798 (45.64) | <0.001 |
| Han ethnicity, n (%) | 14,027 (81.39) | 9,432 (84.94) | 4,595 (74.95) | <0.001 |
| Urban residence, n (%) | 7,661 (44.45) | 5,253 (47.31) | 2,408 (39.28) | <0.001 |
| BMI, kg/m<sup>2</sup>, mean (SD) | 18.38 (3.95) | 19.00 (3.82) | 17.25 (3.95) | <0.001 |
| Near work, h/day, mean (SD) | 4.66 (1.29) | 4.91 (1.25) | 4.22 (1.25) | <0.001 |
| Outdoor activity, h/day, mean (SD) | 3.30 (0.62) | 3.21 (0.62) | 3.44 (0.61) | <0.001 |
| Parental myopia history=0, n (%) | 7,652 (44.40) | 4,914 (44.25) | 2,738 (44.66) | Overall P=0.257 |
| Parental myopia history=1, n (%) | 6,110 (35.45) | 3,982 (35.86) | 2,128 (34.71) | Overall P=0.257 |
| Parental myopia history=2, n (%) | 3,473 (20.15) | 2,208 (19.88) | 1,265 (20.63) | Overall P=0.257 |
| $SE_{\text{mean}}$ D, mean (SD) | $-1.41$ (1.84) | $-2.27$ (1.68) | $0.15$ (0.78) | <0.001 |

Note: Parental myopia history was coded as 0, neither parent; 1, one parent; and 2, both parents. The P value is the overall group-comparison test.

### Linear versus spline models

As shown in Table 2, after adjustment for behavioral, family, and nonlinear BMI terms, the association between rope skipping and both myopia and SE_mean_ remained nonlinear. Relative to the linear models, the spline models reduced AIC by 8.5 for myopia and 4.5 for SE_mean_, with joint tests of nonlinearity yielding P=0.005 and P=0.041, respectively. The spline models for endurance time did not improve fit: ΔAIC was 1.1 for myopia and 1.7 for SE_mean_, with P=0.100 and P=0.249, respectively.

**Table 2.** Comparison of linear and restricted cubic spline models.

| Exposure | Subgroup | Outcome | $\Delta AIC$ | P for nonlinearity |
| --- | --- | --- | --- | --- |
| Rope skipping | 6–12 years | Myopia | −8.5 | 0.005 |
| Rope skipping | 6–12 years | $SE_{mean}$ | −4.5 | 0.041 |
| Endurance time | 16–18 years | Myopia | +1.1 | 0.100 |
| Endurance time | 16–18 years | $SE_{mean}$ | +1.7 | 0.249 |

Note: All models adjusted for age, sex, ethnicity, urban–rural residence, near-work duration, outdoor activity time, parental myopia history, BMI, and centered BMI squared. A negative $\Delta AIC$ indicates improved fit of the spline model.

**Table 3.** Main results of threshold segmented regression.

| Exposure and outcome | Threshold | Effect before threshold | P value | Effect after threshold |
| --- | --- | --- | --- | --- |
| Rope skipping, myopia (OR per 10 skips/min) | 66 | 0.949 (0.919–0.980) | 0.001 | 1.037 (0.999–1.077) |
| Rope skipping, $SE_{mean}$ (D per 10 skips/min) | 66 | +0.034 (+0.016 to +0.052) | <0.001 | −0.014 (−0.042 to +0.014) |
| Endurance time, myopia (OR per 10 s slower) | 307 | 1.020 (0.982–1.058) | 0.310 | 1.255 (1.030–1.531) |
| Endurance time, $SE_{mean}$ (D per 10 s slower) | 307 | −0.027 (−0.066 to +0.012) | 0.180 | −0.098 (−0.171 to −0.026) |
| Rope skipping, myopia (OR per 10 skips/min) | 66 | 0.949 (0.919–0.980) | 0.001 | 1.037 (0.999–1.077) |

Note: Effects are scaled per 10 additional skips/min for rope skipping and per 10 s of slower endurance time. Confidence intervals and P values are based on school-clustered robust standard errors.

**Table 4.** Comparison of key effects after adding waist-to-height ratio to the model.

| Key effect | Primary model: BMI + BMI<sup>2</sup> | Sensitivity model: BMI + BMI<sup>2</sup> + WHtR | Effect unit |
| --- | --- | --- | --- |
| Rope skipping before threshold, myopia | 0.949 (0.919–0.980) | 0.949 (0.919–0.980) | Per 10 skips/min |
| Rope skipping after threshold, myopia | 1.037 (1.000–1.076) | 1.038 (1.001–1.076) | Per 10 skips/min |
| Rope skipping before threshold, $SE_{\text{mean}}$ | +0.034 (+0.016 to +0.052) | +0.034 (+0.016 to +0.052) | Per 10 skips/min |
| Endurance after 307 s, myopia | 1.255 (1.030–1.531) | 1.257 (1.033–1.530) | Per 10 s slower |
| Endurance after 307 s, $SE_{\text{mean}}$ | -0.098 (-0.171 to -0.026) | -0.101 (-0.172 to -0.030) | Per 10 s slower |

Note: WHtR, waist-to-height ratio. WHtR was used as a proxy for adiposity distribution and does not replace direct body-fat or muscle-mass measurements.

### Threshold identification and segmented effects

The AIC grid search identified 66 skips/min as the reference threshold for rope skipping. Below or at this threshold, each additional 10 skips/min was associated with lower odds of myopia (OR 0.949, 95% CI 0.919–0.980; P=0.001) and a 0.034-D shift toward more hyperopic SE_mean_ (95% CI 0.016–0.052; P<0.001). Above the threshold, the protective association did not continue (myopia OR 1.037, 95% CI 1.000–1.076; P=0.048; SE_mean_ −0.014 D, 95% CI −0.042 to 0.014; P=0.316).

For endurance time, the reference threshold was 307 s. Neither outcome showed a statistically significant association before the threshold. Beyond 307 s, each additional 10 s of slower performance was associated with higher odds of myopia (OR 1.255, 95% CI 1.030–1.531; P=0.025) and a 0.098-D shift toward myopia in SE_mean_ (95% CI −0.171 to −0.026; P=0.007). Tests of slope change were not statistically significant for endurance time (P=0.063 for myopia and P=0.092 for SE_mean_), so 307 s is better interpreted as an exploratory tail-risk marker than as a validated biological breakpoint.

### Sensitivity analysis for body-size indicators

After adding WHtR, the directions of the key effects were unchanged. For rope skipping, the threshold-before and threshold-after myopia estimates were OR 0.949 and 1.038, respectively, and the corresponding SE_mean_ estimates were +0.034 D and −0.014 D. Beyond 307 s, the endurance estimates were OR 1.257 for myopia and −0.101 D for SE_mean_. Because WHtR captures only part of abdominal adiposity, it cannot replace direct body-fat or muscle-mass measurements and should not be interpreted as full body-composition adjustment.

## Discussion

### Main findings and interpretation

The reanalysis retained the most consistent structural findings from the original study. Among primary-school students, the association between rope skipping and refractive outcomes was concentrated in the lower-performance range. Among older adolescents, the clearest signal occurred in the slower endurance tail. After adding near-work duration, outdoor activity time, parental myopia history, and a centered BMI-squared term, the direction and magnitude of the main associations changed little. These covariates therefore did not fully account for the observed range-specific patterns^[11–21]^. Nevertheless, the cross-sectional design cannot establish whether physical fitness affects refractive development, whether refractive status affects physical performance, or whether both are influenced by unmeasured factors.

### Body-size confounding and the limits of available measures

The BMI-squared term and WHtR sensitivity analysis partially address the reviewer’s concern, but they cannot substitute for direct body-composition measurements. BMI reflects fat mass, lean mass, and growth-related body size, while WHtR captures only part of fat distribution. The dataset did not include bioelectrical impedance analysis, dual-energy X-ray absorptiometry, or other direct measurements of body composition. We therefore describe the findings as adjusted associations under the available covariate set rather than independent or causal effects.

### Cycloplegic refraction and measurement quality

The study used cycloplegic autorefraction, which reduces the influence of accommodation on refractive measurements in children. For reproducibility, the final submission should specify the cycloplegic agent and concentration, number of instillations, waiting time, bilateral measurement procedure, instrument model, and quality-control procedures.

### Implications for school-based prevention

The 66-skips/min and 307-s thresholds should be used as exploratory prompts for further assessment rather than as pass–fail standards or diagnostic boundaries. Schools could combine fitness results with near-work behavior, outdoor activity, and family history when prioritizing follow-up. A single fitness test should not be used to label an individual student as having a high risk of myopia. These prevention-related considerations are consistent with the wider evidence on outdoor time, screen exposure, sleep, and near work^[22–24]^.

### Limitations and future research

First, the cross-sectional design and convenience sampling limit causal inference and generalizability. Second, direct body-fat percentage, skeletal-muscle mass, axial length, sleep, screen exposure, and socioeconomic variables were unavailable, leaving potential residual confounding^[25–28]^. Third, near-work duration, outdoor activity time, and parental myopia history were obtained from an organized survey dataset; the questionnaire version, recall window, and quality-control procedures should be documented in the final submission. Fourth, although cycloplegic autorefraction was used, the cycloplegic protocol and instrument quality-control details should be reported fully. Future studies should add bioelectrical impedance or DXA measurements, age- and sex-standardized behavioral measures or accelerometry, verified parental refractive status, axial length, sleep and screen exposure, and longitudinal follow-up to test whether the proposed thresholds replicate over time.

## Conclusions

Among Han and Li students in Hainan Province, myopia prevalence was high. After adjustment for age, sex, ethnicity, urban–rural residence, near-work duration, outdoor activity time, parental myopia history, and nonlinear BMI terms, rope skipping in the lower-performance range was associated with lower myopia odds and a less myopic SE_mean_, whereas endurance times beyond 307 s were associated with higher myopia odds and a more myopic SE_mean_. The 66-skips/min and 307-s thresholds may support exploratory school-level risk stratification, but they should not be interpreted as causal intervention targets or individual diagnostic cutoffs. Missing direct body-composition measures and the cross-sectional design limit causal interpretation.

## Data Availability

The data used in this study contain individual-level personal and health-related information from Chinese school students, including minors. In accordance with applicable Chinese requirements concerning national data security, personal-information protection, and research ethics, the raw individual-level dataset cannot be publicly released or deposited in an unrestricted repository. Qualified researchers may submit a reasonable request to the corresponding author at. Requests will be reviewed case by case and will require approval by the data custodian and, where necessary, the relevant ethics review authority. Subject to these approvals, appropriately de-identified aggregate data, a data dictionary, and analysis code may be provided. The provenance and governance of the data are documented by the National Social Science Fund of China, 2025 General Project in Education (grant no. BLA250195), and the Ethics Review Committee of Hainan Normal University (reference no. HNU-260407003). These records provide documentary evidence of the study's funding provenance and ethical oversight but do not authorize public disclosure of protected personal information.

## Data availability

The dataset used in this study contains extensive individual-level physical-fitness, demographic, and refractive data from Chinese children and adolescents. Because the dataset involves minors and potentially sensitive health information, the raw data and any derived data that could permit direct or indirect identification cannot be publicly released or deposited in an unrestricted public repository under applicable national data-security, personal-information protection, and research-ethics requirements. Accordingly, the research team does not provide open download access.

Journal editors and reviewers who require access for peer review may submit a reasonable request to the corresponding author, describing the purpose of verification and the proposed data-security arrangements. Subject to approval by the data custodian and, where necessary, the relevant ethics oversight body, the research team may provide de-identified aggregate data, analysis code, a data dictionary, or other restricted-access materials sufficient to verify the main findings.

The provenance and regulatory compliance of the dataset can be supported by the National Social Science Fund project approval documentation, the ethics approval issued by the Social Sciences Ethics Committee of Hainan Normal University, and related survey and data-management records. All data-access requests will be reviewed on a case-by-case basis in accordance with applicable data-security and research-ethics requirements.

## Acknowledgments

The authors thank the participating schools, students, guardians, physical-education researchers, and optometry staff for their cooperation in data collection.

## Author Contributions

Xin Li: Data curation and formal analysis.

Aichun Li: Physical-fitness testing and field data collection.

Junlai Zhou: Project administration and coordination with participating schools and relevant departments. All authors reviewed and approved the final manuscript.

